# Chyme Reinfusion Practices in the Neonatal Population

**DOI:** 10.1101/2024.09.06.24312922

**Authors:** Alexandria H Lim, Georges Tinawi, Taylor Harrington, Emma Ludlow, Helen Evans, Ian Bissett, Celia Keane

**Affiliations:** Department of Surgery, The University of Auckland, Auckland, New Zealand; Department of Paediatric Surgery, Wellington Hospital, Wellington, New Zealand; Starship Children’s Hospital, Te Toka Tumai Auckland, Te Whatu Ora, New Zealand; The Insides Company, New Zealand; Department of Surgery, Te Toka Tumai Auckland, Te Whatu Ora, New Zealand; Department of Surgery, Te Tai Tokerau, Te Whatu Ora, New Zealand

## Abstract

**Introduction:** The utilisation of chyme reinfusion therapy (CRT) by returning the output from the proximal limb into the distal limb of double enterostomies is a safe and effective method to improve nutritional uptake and maintain intestinal integrity in adult populations. This technique is also particularly suitable for neonatal patients with life-threatening conditions such as congenital bowel abnormalities and necrotising enterocolitis (NEC). Despite its promise, it has only had irregular uptake in neonatal patients. We aimed to identify the frequency, methodology and adverse events associated with CRT in the appropriate neonatal population.

**Methods:** A ten-year retrospective cohort study was conducted using database searches at two major paediatric hospitals in New Zealand. All patients with suitable anatomy were identified, and data on CRT methodology and outcomes were recorded.

**Results:** Of the 49 neonates identified with double enterostomy 23 (47%) underwent CRT for high stoma output, risk of short gut syndrome or as a routine protocol before re-anastomosis. A nasogastric feeding tube was inserted into the distal limb and collected chyme was reinfused via manual bolus or automated syringe driver. The median (IQR) weight gain increased from 13.9 (3.50 - 22.89) to 24.37 (19.68 - 29.99) g/day during CRT (p = 0.04). 18 infections requiring medical intervention but unrelated to CRT occurred in 13 patients (56%). The rate of non-infectious adverse events was 24, with 7% remaining free from any adverse events.

**Conclusion:** Chyme reinfusion is an underutilised method of improving nutrition in neonates with intestinal failure. Premature neonates requiring double enterostomy formation are at high risk of infectious and non-infectious complications, but few of these are related to CRT. Standardised protocols providing clear eligibility criteria and detailed methodology for CRT are required to promote uniform utilisation of this practice.

## Introduction

Intestinal failure is a highly morbid diagnosis characterised by digestive and absorptive deficits secondary to loss of functional gut length requiring parenteral nutrition. Neonates are particularly susceptible to intestinal failure in the setting of necrotising enterocolitis, malrotation with volvulus and other congenital conditions that result in the loss of small bowel length. These conditions often require the formation of an enterostomy during the neonatal period. Parenteral nutrition is required in these neonates but is associated with high rates of central line infections and long-term hepatobiliary complications (1).

Chyme reinfusion (CR) mimics the state of intestinal continuity providing valuable nutritional support for patients with intestinal failure, reducing intravenous fluid, electrolyte, and nutritional requirements and promoting distal intestinal function (2). It is also a promising pre-operative strategy to promote intestinal rehabilitation. The safety and feasibility of chyme reinfusion in adults is more readily established and acceptable within the literature (2–6); however, the use of chyme reinfusion in neonatal populations remains limited (7–9). Additionally, there is a paucity of published protocols to guide standardised methods of chyme reinfusion in neonates.

We therefore aimed to describe the practice of distal chyme reinfusion in neonates and children in two tertiary institutions in New Zealand. Our primary objective was to determine the rate of gastrointestinal infection post chyme reinfusion in this neonatal cohort, and also to identify the methodology and practice of chyme reinfusion and evaluate the wider morbidity, mortality, and outcomes of chyme reinfusion.

## Methods

This was a retrospective cohort study from two tertiary centres in New Zealand; Starship Children’s Hospital (Auckland) and Wellington Hospital. Ethical approval was granted by the Health and Disability Ethics Committee (HDEC) EXP14061, with locality approval at both sites (2023 LOCAL 14061 and 2023-CCHV-61).

### Study Population

Patients aged < 18 years who underwent chyme reinfusion between December 2012 and December 2022, for any indication, were eligible to participate. Eligible participants were identified retrospectively via the Neonatal Intensive Care Unit (NICU) and Paediatric Surgery databases and cross-matched with clinical coding data (see **Figure 1**). Paper and electronic medical records were screened to identify potentially eligible patients with either double enterostomy (DES) or entero-atmospheric fistula (EAF). These were then screened further to determine those that had undergone refeeding during the study period. All methods of chyme reinfusion were considered eligible including manual and automated reinfusion, as well as bolus and continuous reinfusion methods.

**Figure 1.**
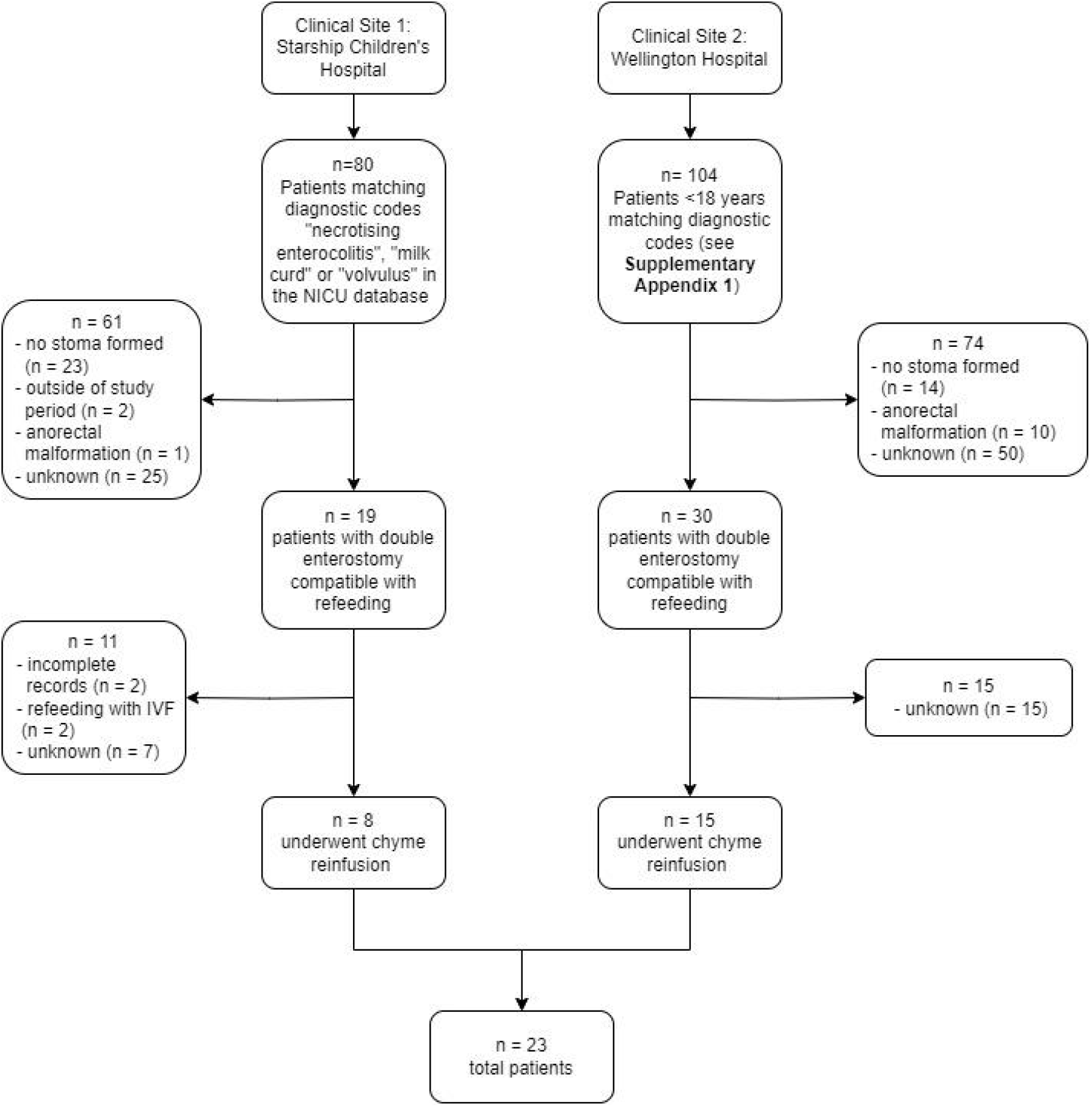
Participant Flow.

Starship Children’s Hospital performed refeeding upon the recommendation of multidisciplinary input (including paediatric doctors, gastroenterologists, surgeons and dieticians), primarily led by the surgical team. Although there were no formal criteria, chyme reinfusion was considered for patients with high stoma outputs and significant comorbidities, such as faltering growth crossing major percentile lines or liver dysfunction secondary to total parenteral nutrition (TPN). The decision to initiate refeeding at Wellington Hospital was predominantly surgeon-dependent, with a wide variation in individual practice. The surgical team determined reinfusion rates and volumes and monitored for complications. Passing stool per rectum was considered to be the definition of successful refeeding. In the latter study period there was increased uptake of chyme reinfusion within the paediatric surgical department but this was not protocolised at an institutional level.

### Data Collection

Demographics and clinical characteristics, including gestational age, sex, ethnicity, weight, primary pathology, and stoma characteristics were extracted. The indications for stoma formation, chyme reinfusion and cessation of chyme reinfusion were collected. The following reinfusion characteristics were extracted: stoma output, volume reinfused, duration of reinfusion, health professional performing reinfusion, and method of administration. The incidence and severity (Clavien-Dindo classification) of documented complications were recorded as were the outcomes (nutrition, growth, post-restoration of bowel continuity) of chyme reinfusion. Nutritional outcomes were assessed as an overall change in feeding method during the reinfusion period. Growth was assessed using change in weight per day (g/day) according to the following formulae:

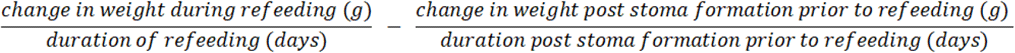

Change in weight during refeeding was calculated by deducting the patient’s weight on the day refeeding was commenced from the weight at the end of the refeeding period. Change in weight post stoma formation prior to refeeding was calculated by deducting the patient’s weight on the day of stoma formation from the weight on the day refeeding was commenced.

### Statistical Analysis

Data was primarily reported using descriptive statistics. Parametric data are reported as mean and standard deviation. Non-parametric data are presented as the median and interquartile range (IQR) or range. Spearman’s correlation and multiple linear regression models were used to assess the association between clinical and reinfusion variables and weight-related outcomes. Correlations were considered weak if below 0.4, moderate between 0.4 and 0.78 and strong if they were greater than 0.8. Significance was determined if p<0.05. Analyses were performed using R software (Version 4.2.3, R Foundation for Statistical Computing, Vienna, Austria).

## Results

There were 184 participants identified from database and clinical coding searches. By identifying those with a double enterostomy within the study period, the sample was reduced to 49 patients who may have undergone refeeding. A total of 25 patients underwent chyme reinfusion over the study period, but 2 were lost to follow-up and excluded due to transfer to other hospitals. The final study population totalled 23 neonates (see **Figure 1**). **Table 1** outlines the participant characteristics including primary pathology and indication for stoma formation. A full summary of individual patient characteristics, stoma characteristics and refeeding methods is displayed in **Supplementary Table 1** and a full list of diagnostic codes used at Wellington Hospital is available in **Supplementary Appendix 1**.

**Table 1.** Participant Characteristics.

| Characteristic |  |
| --- | --- |
| Median (IQR) gestational age at birth (weeks) | 27 (25 - 28) |
| Median (IQR) gestational age at stoma formation (weeks) | 30 (27.5 - 35.5) |
| Female, n (%) | 14 (61%) |
| Ethnicity, n (%)<br>- NZ European<br>- Maori<br>- Samoan<br>- Other European<br>- Tongan<br>- Indian<br>- African | 9 (39%)<br>5 (22%)<br>3 (13%)<br>3 (13%)<br>1 (4%)<br>1 (4%)<br>1 (4%) |
| Indication for stoma formation<br>- Necrotising Enterocolitis (NEC)<br>- Meconium ileus (including associated ileal atresia n = 1)<br>- Volvulus<br>- Milk curd obstruction<br>- Perforation cause not otherwise specified<br>- Anastomotic leak | 8 (35%)<br>5 (22%)<br>3 (13%)<br>1 (4%)<br>4 (17%)<br>2 (9%) |
| Median (IQR) length of bowel resected (cm) * | 10 (0 - 100) |
| Median length of small bowel remaining, cm (range)** | 60 (28 - 145) |
| Stoma<br>- Ileostomy<br>- Ileal distal limb<br>- Caecal mucous fistula<br>- Ascending colon mucous fistula<br>- Transverse colon mucous fistula<br>- Jejunostomy<br>- Jejunal distal limb<br>- Ileal mucous fistula<br>- Colonic mucous fistula | 12<br>6<br>2<br>4<br>2<br>11<br>2<br>8<br>1 |
| Median weight at stoma formation, g (range) | 1217 (700 - 4110) |

\*data unavailable from 4 participants
\*\*data only available for 9 participants

### Chyme Reinfusion

Chyme reinfusion was routinely carried out by the neonatal nurses at the patient’s bedside for all 23 participants. At Starship Children’s Hospital, 6 participants used a specifically designed refeeding system, The Insides^®^ Neo (see **Figure 2**). The remaining two participants from this centre had chyme collected from their stoma bags at 4-6 hour intervals and reinfused via a size 6 or 8Fr gastric tube into the distal limb, with a small incision made in the top of the bag to allow insertion of the tube into the distal lumen, secured with medical tape, and attached to a syringe driver. Reinfusion occurred continuously over a 4-6 hour period at a variable rate to minimise reflux from the distal limb and any additional volume unable to be infused was discarded. Reinfusion was carried out at a median of four times daily and adjusted based on the volume of stoma output. At Wellington Hospital all but one participant had manual bolus reinfusion using a syringe via a 5 or 6 Fr nasogastric tube over a period of several minutes. If there were concerns with reflux of chyme during refeeding, a Foley catheter was used instead of a nasogastric feeding tube with the balloon of the Foley inflated externally to provide a seal across the distal limb. The frequency of refeeding was increased as tolerated so that by the end of the refeeding period, neonates were refed 1-8 times daily with the largest number being refed four times daily. Reinfusion-related metrics are reported in **Table 2**.

**Table 2.**
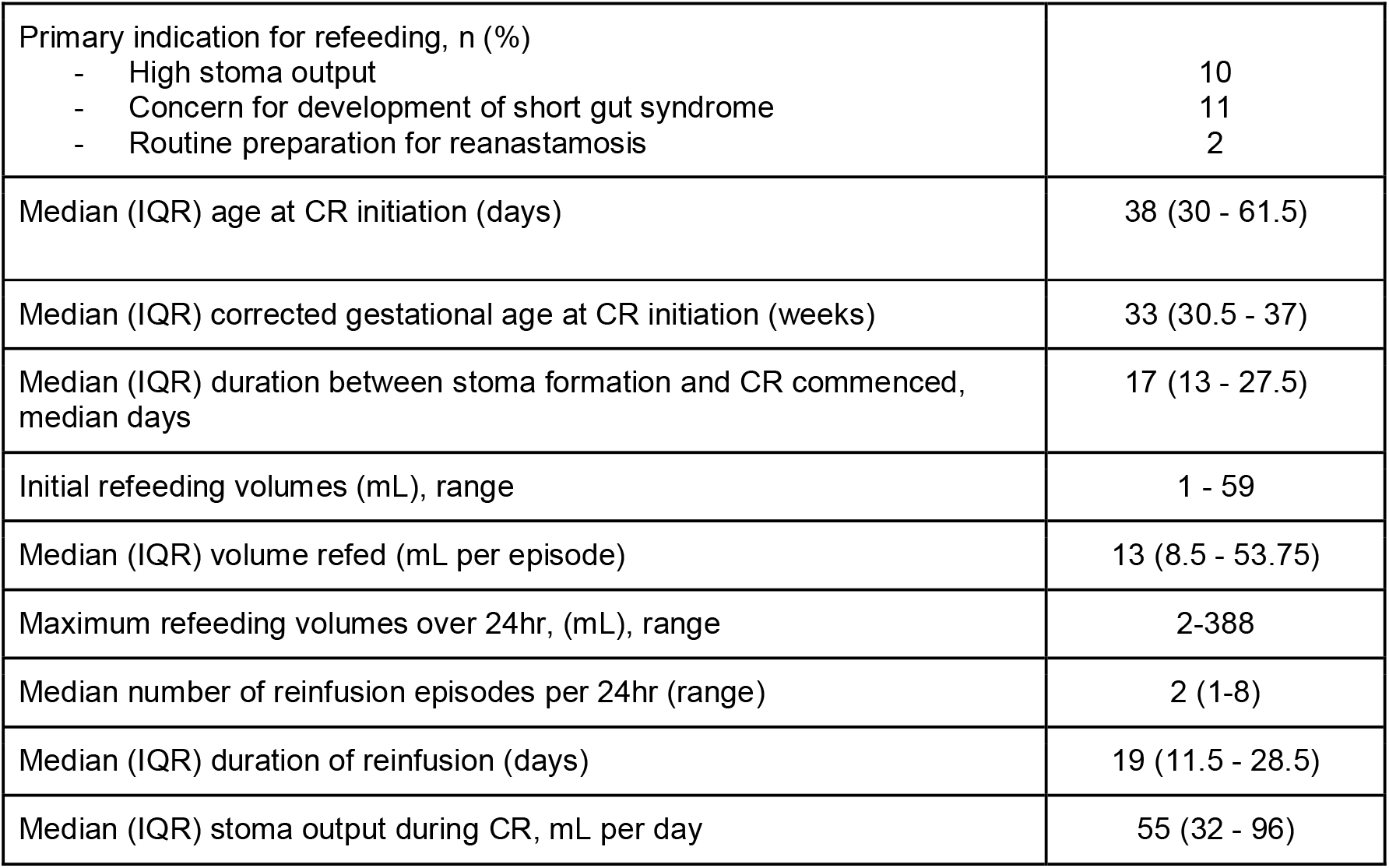
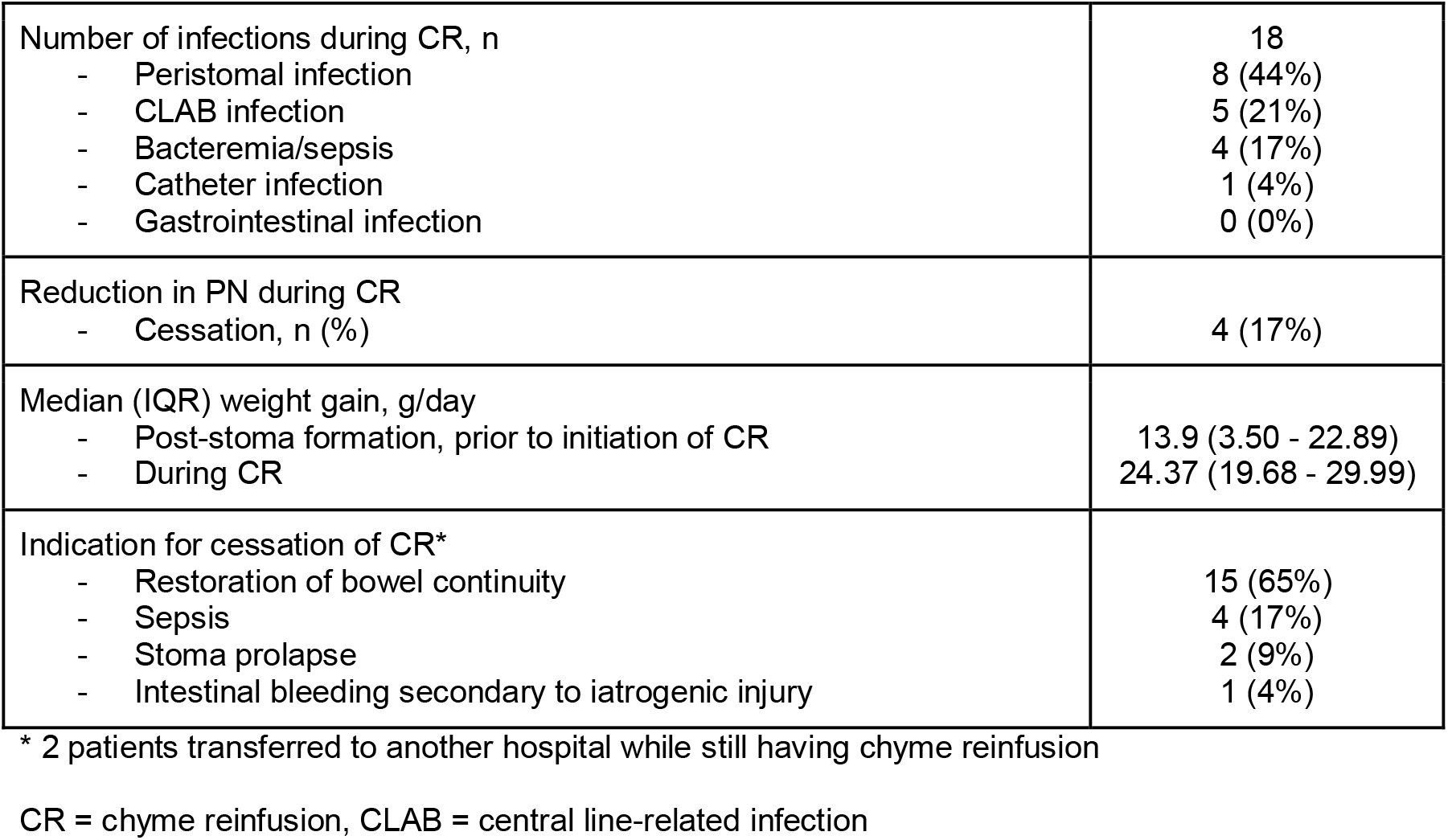
Chyme Reinfusion Characteristics and Outcomes.

**Figure 2:**
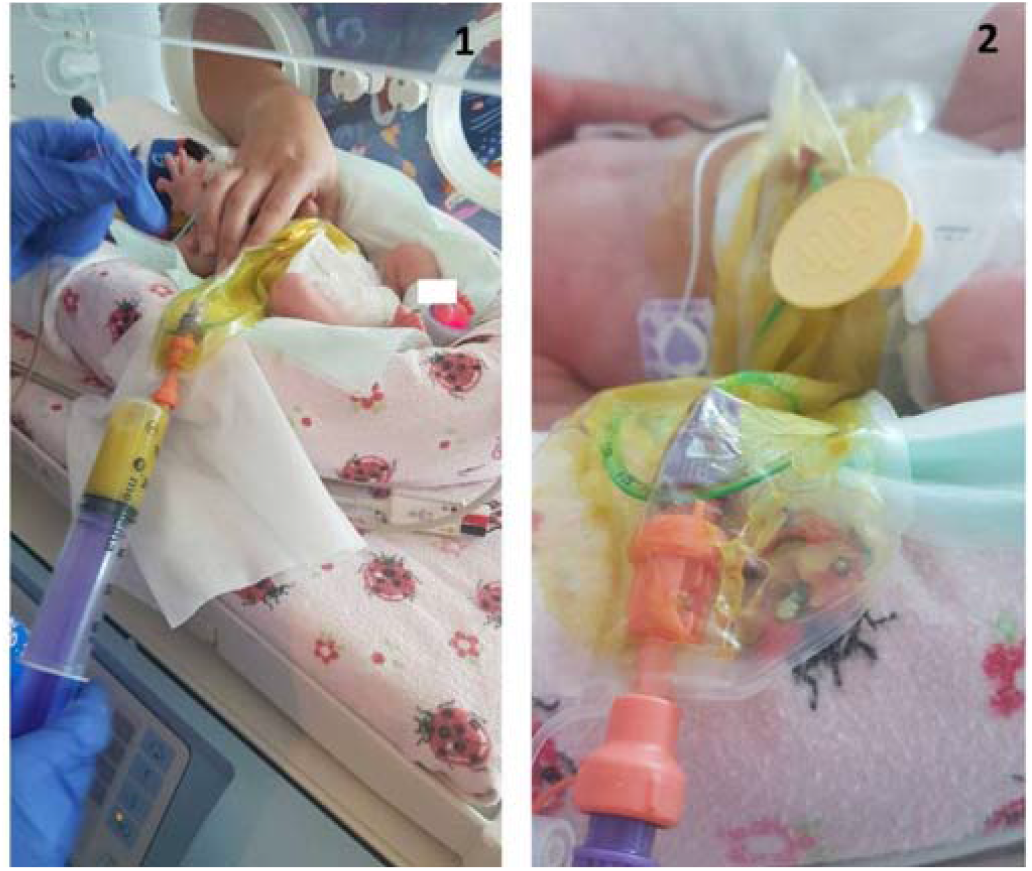
The neonatal chyme reinfusion system. 1 = Syringe attachment and aspiration of chyme in preparation for reinfusion, 2 = Housing component attached to gastric feeding tube facilitating delivery of chyme to distal limb. Published with parental permission.

### Complications

During and after the period of chyme reinfusion, there were no infectious complications that could be directly attributed to reinfusion. However, this population was at high risk of morbidity, and 13 (56%) patients suffered from infections unrelated to chyme reinfusion that required medical intervention.

There were seven incidents of peristomal skin infection requiring antibiotics, five incidents of central-line associated bloodstream (CLAB) infection, four incidents of sepsis where the source was not clear, one stoma wound abscess and one urinary catheter infection. The vast majority of complications were managed at Clavien-Dindo Grade II, with the exception of the stoma wound abscess requiring incision and drainage under general anaesthetic (Clavien-Dindo Grade IIIa).

There was a high incidence of non-infectious adverse events that could not be directly attributed to chyme reinfusion. The most common adverse event was peristomal skin irritation, occurring in 14 (60%) patients. Six (26%) patients suffered from stomal haemorrhage, with a total of eight haemorrhages occurring during refeeding. There were 11 stomal prolapses in 10 (43%) patients, with one case requiring chyme reinfusion cessation as a result. Additionally, there were 2 parastomal hernias managed conservatively and 1 episode of stoma wound dehiscence. There were 2 participants (7%) who did not experience any complications during the refeeding period.

Two patients were transferred to another hospital before restoration of bowel continuity resulting in 21 (91%) patients achieving restoration of bowel continuity within the study period and being able to be assessed for post-stoma reversal outcomes. Post-operatively, there were three occurrences of CLAB, a single occurrence of each of; surgical wound infection, post-operative pneumothorax, incisional hernia as a result of re-anastomosis and bowel obstruction.

### Other outcomes

Eight participants (35%) who underwent chyme reinfusion reduced or ceased parenteral nutrition. Of these patients, three (38%) were able to increase enteral nutrition as a result. There were 2 patients (9%) who initiated TPN during the refeeding period, once to maintain sodium intake on a background of cystic fibrosis, and one for reasons unknown. Patients gained a median of 14.3g extra per day during refeeding compared to the period prior to commencing refeeding, resulting in an additional 271g weight gain during the refeeding period based on a median refeeding period of 19 days. Weight gain increased during refeeding compared to the period prior to refeeding (13.9, 23.47, p = 0.04). There were no significant relationships between the amount of weight gained or lost during refeeding and study site, age, ethnicity, gestational age at birth, weight at stoma formation, day of life reinfusion commenced, gestational age at the commencement of reinfusion, number of days post-stoma formation at the commencement of reinfusion. There was a strong correlation between younger gestational age at stoma formation and increased weight change over the refeeding period (Spearman’s coefficient of -0.80). The full comparison is outlined in **Supplementary Table 2**. Change in weight prior to refeeding did not have an effect on change in weight during refeeding (p = 0.2) or average weight gained or lost (p = 0.06), whereas there was a strong relationship between weight gained or lost during refeeding and average weight gained or lost (p < 0.001).

## Discussion

This ten-year descriptive study of two tertiary hospitals in NZ only identified 23 paediatric patients receiving chyme reinfusion. Chyme reinfusion accelerated weight gain and promoted increased nutritional intake for neonates with double enterostomies. Neonates were shown to gain additional weight during the period of chyme reinfusion regardless of when refeeding was commenced and the degree of stoma output without evidence of increased rates of adverse events and structural complications. At two major clinical sites in a developed country, less than 50% of eligible neonates are undergoing chyme reinfusion, indicating there are significant barriers to its use. These will need to be identified and overcome to improve the implementation of chyme reinfusion practices in routine clinical practice.

Neonates with intestinal failure and stoma formation have high rates of adverse events and complications requiring treatment in up to 80.6% of patients with all aetiologies considered (10). There was a high rate of clinical complications requiring intervention that could not be directly attributed to chyme reinfusion. Infectious complications requiring intervention occurred at 78% (n = 18 infections in 23 patients) in our population, a low rate in comparison to earlier neonatal intestinal failure populations who had not undergone refeeding (10–12). The rates of infection and complication may be attributable to the frailty of the patient population and the requirement of high-intensity intervention such as TPN and long-line insertion, rather than refeeding itself, as the lowest reported rate is still high at 34% (13). Peristomal skin irritation, the most common contributor, often occurred in isolation in our study without additional intervention or complications. Stoma complications during refeeding were successfully managed with minimally invasive therapeutic strategies, and surgical intervention occurred at a reduced rate than other populations of neonates with stomas (10). Adverse events did not prevent stoma closure in any instance. After the formation of a stoma, refeeding did not result in an increase in stoma complications such as stomal prolapse or dysfunction compared to other studies with similar populations (12,14). A lower rate of complications post-stoma formation suggests chyme reinfusion may be a safe method to use in the neonatal population and may play a role in decreasing morbidity due to the benefits of additional nutritional support and weight gain.

Sufficient nourishment is difficult to achieve in neonates with high-output stomas and intestinal failure (12,15). This study has demonstrated an improvement in weight gain after refeeding in keeping with the most recent literature, supporting the beneficial role of neonatal chyme reinfusion to promote increased nutritional intake (16,17). Barker hypothesised that nutritional uptake during the neonatal period affects a child’s susceptibility to chronic disease and metabolic capabilities in later life. This makes appropriate nutrition in the neonatal period paramount to ensuring continued health during childhood and the development of a healthy adult body (18). The observed reduction in TPN requirements, increased nutritional intake and eventual TPN cessation in some neonates is promising. This should reduce the number of TPN-related infections and co-morbidities faced by these patients and also promote ongoing health as the child ages. Increased weight gain was demonstrated in all the groups of neonates, suggesting that it should be considered in all neonates with high stoma output and access to distal refeeding.

This is the first Australasian study to demonstrate the safety of chyme reinfusion in neonates. Chyme reinfusion is becoming increasingly recognised as a solution for high-output enterostomy in children (19,20), underlining the importance of reporting the strategies and outcomes of centres currently performing chyme reinfusion. We present two different methods for chyme reinfusion in urban centres that support a nurse-led, multi-disciplinary team approach. The retrospective nature of the study precluded an analysis of the factors influencing decision making and in particular a surgeon’s decision to consider refeeding for a particular neonate. The lack of institutionalised protocols at either institution likely resulted in an inherent selection bias (with various factors being considered at either hospital) as to which neonates were offered refeeding. As such, we recognise the heterogeneity of our cohort; however, given the paucity of data from Australasia regarding neonates undergoing refeeding, we believe our data are a valuable insight into real-world practice at two tertiary institutions in New Zealand. Chyme reinfusion remains underutilized and increased implementation should be actively supported.

This study underscores chyme reinfusion improves growth in premature neonates with double enterostomies, yet it appears to be underutilizsd within the neonatal population. Weight gain during the first few months of life is crucial for ongoing organ development and compensatory growth, serving as a well-established predictor of neurological and retinal development (21–23). During the refeeding period, weight gain improved, nearly reaching the expected levels for infants aged 0-3 months in New Zealand (24). However, over 50% of eligible neonates were not refed, likely due to previously discussed reasons, indicating that the clinical utility of chyme reinfusion has substantial room for expansion. Further research is necessary to establish clear eligibility criteria for refeeding, identify characteristics that lead to favourable growth outcomes, and evaluate the long-term growth patterns of these neonates into childhood.

In conclusion, chyme reinfusion appears to be underutilised but safe for neonates with intestinal failure and an accessible enterostomy distal limb. In such a vulnerable population, with extreme risk of morbidity, a therapy that may have great benefit needs routine consideration rather than occasional contemplation. Paediatric units should be encouraged to standardise their eligibility criteria and methodology for chyme reinfusion so that all appropriate patients have access to this beneficial therapy. The next step is to prospectively evaluate outcomes in larger populations using standard protocols. Ideally, this would include comparative populations, and report longer-term nutritional and developmental outcomes as well as patient and family-centre outcomes such as quality of life.

## Data Availability

Data used for analysis will be made available upon reasonable request, conditional on ethical approvals

https://doi.org/10.6084/m9.figshare.26892595.v1

## Supplementary Material

**Supplementary Table 1:** Table 1 demonstrates the patient characteristics, refeeding regime and outcomes for the neonatal population that underwent chyme reinfusion at two major clinical paediatric hospitals. Available at https://doi.org/10.6084/m9.figshare.26892595.v1

**Supplementary Appendix 1:**
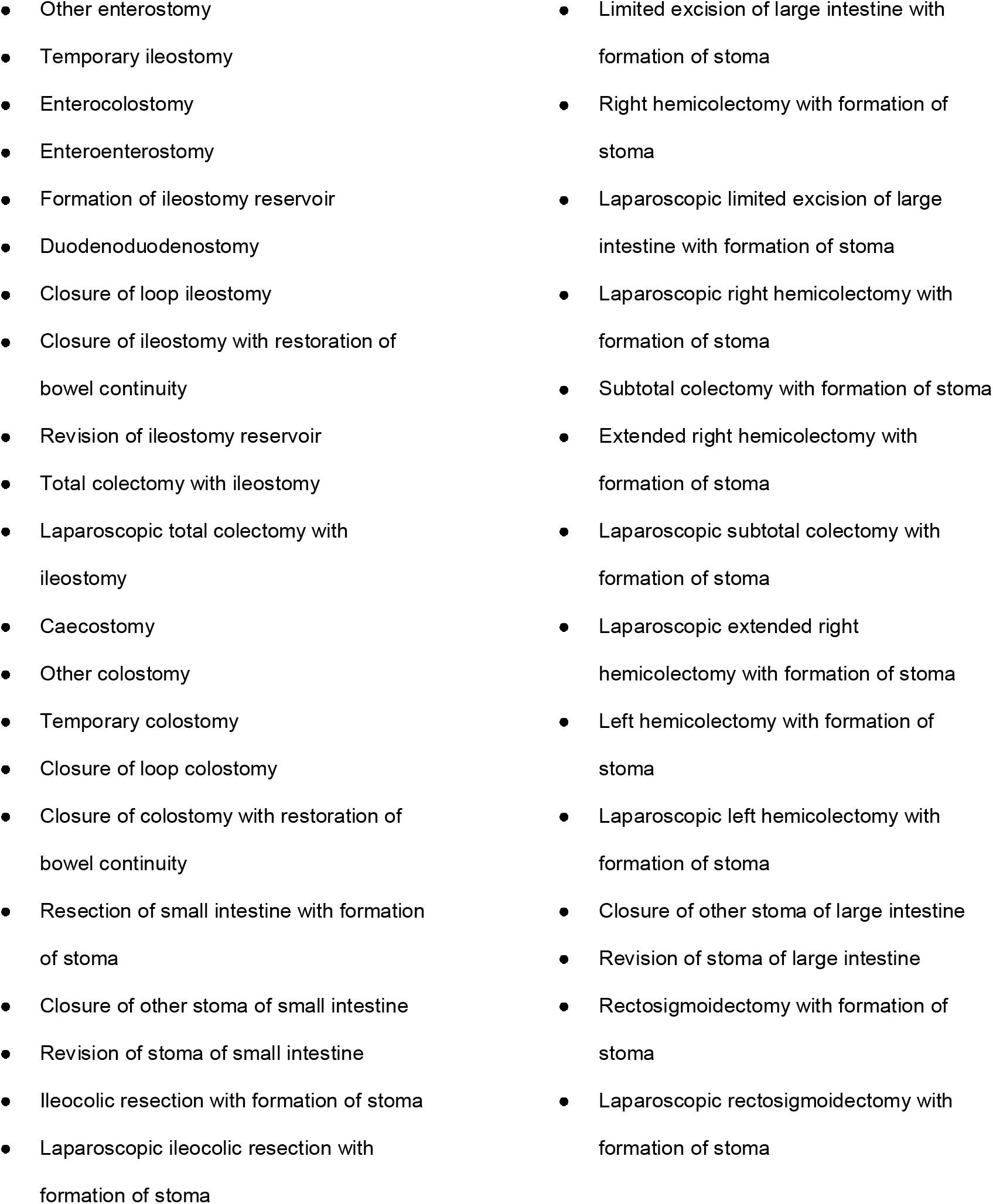
Diagnostic codes used at Wellington Hospital.

**Supplementary Table 2:** Spearman’s Coefficient correlations in reference to average weight change over the refeeding period.

| Variable | Spearman's Coefficient |
| --- | --- |
| Gestational age at birth | 0.30 |
| Gestational age at stoma formation | -0.80 |
| Weight at stoma formation | 0.26 |
| Day of life refeeding commenced | -0.36 |
| Gestational age at commencement of refeeding | 0.07 |
| Number of days post-stoma formation at commencement of refeeding | 0.07 |

